# Radiomic analysis of patient and inter-organ heterogeneity in response to immunotherapies and BRAF targeted therapy in metastatic melanoma

**DOI:** 10.1101/2024.04.26.24306411

**Authors:** Alexandra Tompkins, Zane N. Gray, Rebekah E. Dadey, Serafettin Zenkin, Nasim Batavani, Sarah Newman, Afsaneh Amouzegar, Murat Ak, Nursima Ak, Taha Yasin Pak, Vishal Peddagangireddy, Priyadarshini Mamindla, Sarah Behr, Amy Goodman, Darcy L. Ploucha, John M. Kirkwood, Hassane M. Zarour, Yana G. Najjar, Diwakar Davar, Rivka Colen, Jason J. Luke, Riyue Bao

## Abstract

**Background:** Variability in treatment response may be attributable to organ-level heterogeneity in tumor lesions. Radiomic analysis of medical images can elucidate non-invasive biomarkers of clinical outcome. Organ-specific radiomic comparison across immunotherapies and targeted therapies has not been previously reported.

**Methods:** We queried UPMC Hillman Cancer Center registry for patients with metastatic melanoma (MEL) treated with immune checkpoint inhibitors (ICI) (anti-PD1/CTLA4 [ipilimumab+nivolumab; I+N] or anti-PD1 monotherapy) or BRAF targeted therapy. Best overall response was measured using RECIST v1.1. Lesions were segmented into discrete volume-of-interest with 400 radiomics features extracted. Overall and organ-specific machine-learning models were constructed to predict disease control (DC) *versus* progressive disease (PD) using XGBoost.

**Results:** 291 MEL patients were identified, including 242 ICI (91 I+N, 151 PD1) and 49 BRAF. 667 metastases were analyzed, including 541 ICI (236 I+N, 305 PD1) and 126 BRAF. Across cohorts, baseline demographics included 39-47% female, 24-29% M1C, 24-46% M1D, and 61-80% with elevated LDH. Among patients experiencing DC, the organs with the greatest reduction were liver (−88%±12%, I+N; mean±S.E.M.) and lung (−72%±8%, I+N). For patients with multiple same-organ target lesions, the highest inter-lesion heterogeneity was observed in brain among patients who received ICI while no intra-organ heterogeneity was observed in BRAF. 267 patients were kept for radiomic modeling, including 221 ICI (86 I+N, 135 PD1) and 46 BRAF. Models consisting of optimized radiomic signatures classified DC/PD across I+N (AUC=0.85) and PD1 (0.71) and within individual organ sites (AUC=0.72∼0.94). Integration of clinical variables improved the models’ performance. Comparison of models between treatments and across organ sites suggested mostly non-overlapping DC or PD features. Skewness, kurtosis, and informational measure of correlation (IMC) were among the radiomic features shared between overall response models. Kurtosis and IMC were also utilized by multiple organ-site models.

**Conclusions:** Differential organ-specific response was observed across BRAF and ICI with within organ heterogeneity observed for ICI but not for BRAF. Radiomic features of organ-specific response demonstrated little overlap. Integrating clinical factors with radiomics improves the prediction of disease course outcome and prediction of tumor heterogeneity.

## Background

Despite improvements with BRAF and immune-checkpoint inhibitors (ICI), at least half of patients with melanoma succumb to disease^1 2^. Tissue-based biomarker testing requires invasive procedures and inadequately describes response. Meanwhile, inter-lesion metastatic heterogeneity impacts survival of ICI and is influenced by site of metastasis^3–6^.

Factors intrinsic to tumor cells, such as tumor mutational burden and immunogenicity, among others, vary by anatomic location of metastasis and impact therapy^7–9^. In melanoma and non-small cell lung cancer (NSCLC), metastasis-specific patterns impact the treatment outcome of anti-PD1^3–5^. Inter-lesion heterogeneity of immune cell content has also been described in ovarian and colorectal cancers where non-responding lesions have been associated with immune exclusion^10 11^. Particularly myeloid cells are known to have a complex phenotypic distribution across organs^12^ and impact on antitumor immunity^13 14^. Local therapies for oligometastatic progression to extend BRAF or immunotherapy benefit is a clinical strategy in melanoma oncology^15 16^. An unmet need remains for non-invasive biomarkers of lesion and organ-specific response to inform treatment selection and the study of tumor resistance.

Multiple previous studies have utilized radiomics to predict ICI responses in melanoma, however, these have focused on anti-PD1 or analyzed only single lesions or a predefined set of features to predict patient outcomes^17–21^. Few studies have focused on identifying features specific to metastatic sites. We report patient and organ-level response patterns as well as radiomic models across treatment modalities of ipilimumab plus nivolumab, anti-PD1 monotherapy, and BRAF±MEK inhibitor combinations in advanced melanoma.

## Methods

A full description is supplied in the **Supplementary Methods**. Patients with unresectable stage III/IV melanoma who received ICI (anti-PD1/CTLA4 [nivolumab plus ipilimumab], referred to as I+N cohort, *n*=91; anti-PD1, referred to as PD1 cohort, *n*=151; 242 total), or BRAF±MEKi targeted therapy (BRAF cohort, *n*=49) from 2015-2020 were identified from UPMC Hillman Cancer Center registry (**Fig. S1**; **Table S1**), according to an Institutional Review Board approved protocol (STUDY20020107). Best overall response and organ-specific tumor responses (adrenal, brain, liver, lymph node [LN], lung, and soft tissue) were evaluated by Response Evaluation Criteria in Solid Tumors (RECIST) v1.1. Across all cohorts, 1,166 CT and 168 MRI scans at baseline and best response were analyzed. After quality control, scans were segmented by 3D Slicer, with 400 radiomics features extracted^22^, including 10 first-order (FOF001-010), 195 second-order (SOF001-195), and 195 volume-adjusted second-order features (SOVF001-195). Machine-learning (ML) models by XGBoost were constructed to predict two classes: disease control (DC, including complete response [CR], partial response [PR], stable disease [SD]) or progressive disease (PD). Models were developed to predict overall response or organ-specific response. Two types of ML models (radiomics features only; radiomic plus clinical features) were developed when appropriate. Statistical analysis was performed using R (v4.1.2) and Bioconductor (release 3.14), with false discovery rate (FDR) controlled at 0.10, and Benjamini-Hochberg (BH)-FDR adjustment for multiple comparisons. An analysis overview is provided in **Fig. S2**, with the 400 radiomics features described in **Tables S2** and **S3**.

## Results

### Population Characteristics and Overall Response

Among 242 patients receiving ICI (I+N and PD1), 199 (82%) received first-line immunotherapy (**Table S1**). The I+N and BRAF cohorts had higher combined incidences of M1C (extra-pulmonary metastasis) and M1D (central nervous system metastasis) by American Joint Cancer Classification (AJCC) staging criteria (69% and 70%, respectively), while PD1 showed similar distribution across M1A-D. In all cohorts, the majority of patients exhibited high levels of circulating lactate dehydrogenase (LDH>upper limit normal[ULN], the institutional ULN of LDH is 170) and elevated neutrophil-to-lymphocyte ratio (NLR>3.0) at baseline. DC of 53% (8% CR, 25% PR, 20% SD) was observed in all ICI (**Fig. 1A**). Cohort review included I+N, PD1, and BRAF, respectively, with 41%, 60%, and 86% DC (**Fig. 1A**). DC varied by line of therapy with I+N (45% first line *versus* 31% in ≥2 lines) and PD1 (62% first line *versus* 41% in ≥2 lines; **Fig. 1B**), consistent with the literature^23^.

**Figure 1.**
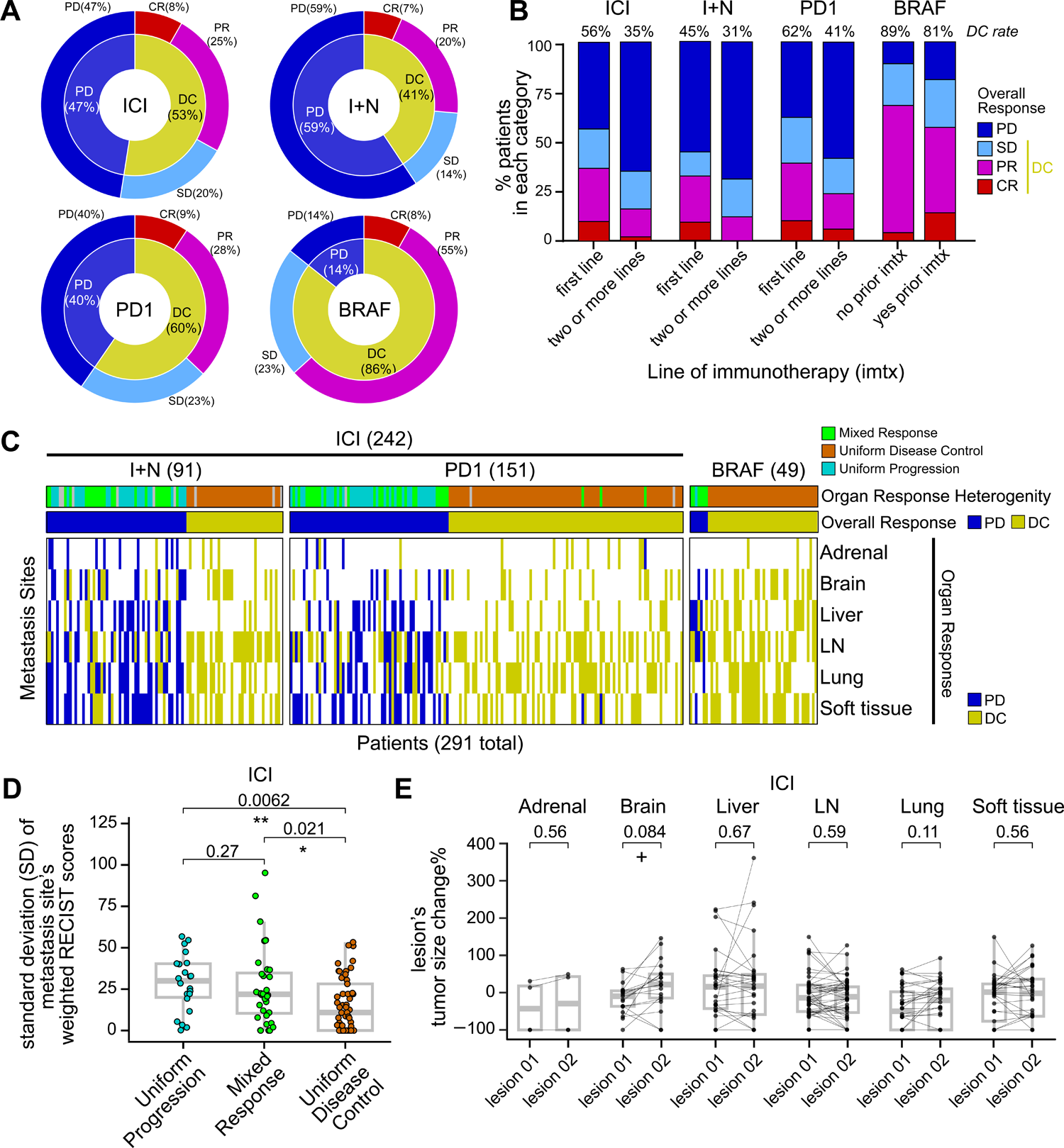
Overall and organ-specific response in ICI (I+N, PD1) and BRAF cohorts. (**A**) Overall response to therapy by treatment. For each cohort, the outer circle shows percentage of patients who experienced CR, PR, SD, or PD. The inner circle shows DC% (including CR, PR, and SD) and PD%. (**B**) Patients stratified by previous exposure to immunotherapy. Color represents CR, PR, SD, and PD same as in **A**. Number above each bar shows the percentage of patients who experienced DC in each subset. Imtx = immunotherapy. (**C**) Heatmap showing the organ-specific response (adrenal, brain, liver, LN, lung, soft tissue, on the row) and in the context of overall response per patient (on the column). *n*=291 patients are shown in **A**, **B**, and **C**, with 91 from I+N, 151 from PD1, and 49 from BRAF. (**D**) Comparison of inter-organ heterogeneity in patients with mixed response *versus* those with uniform progression *versus* those with uniform disease control. The y-axis represents the standard deviation of weighted RECIST scores across all metastases in a patient. Each data point represents one patient. *n*=111 ICI patients who had at least two metastasis sites are shown. (**E**) Intra-organ heterogeneity by organ site comparing lesions 01 and 02. The y-axis represents individual lesion’s tumor size change in percentage. Each data point represents one lesion. Lines connect lesion 01/02 from the same metastasis site in the same patient. *n*=168 sites from ICI patients who had two lesions per site are shown. LN = lymph node. Wilcoxon rank-sum test was used in **D**, Wilcoxon signed-rank test was used in **E**. FDR-adjusted p-values are shown in **D** and **E**. FDR was controlled at 0.10. All tests are two-sided. Denotations: ** *P*<0.01, * *P*<0.05, + *P*<0.10.

### Mixed organ responses are associated with a higher risk of progression at the patient level

Patients with mixed responses between metastatic sites had a significantly higher likelihood of PD, suggesting heterogeneity in organ-specific response determines overall response (*P*<0.0001 in all cohorts, two-sided Fisher’s exact test; **Fig. 1C**). Organ-specific response was defined by weighted RECIST (referred to as, RECIST_weighted_), taking into account the baseline size as well as lesion size change at best response *versus* baseline, if more than one lesion was identified per organ (see **Methods**). Patients were categorized into uniform PD, mixed response if some organs were DC and some were PD, and uniform DC. Patients with uniform PD showed significantly higher variation of RECIST_weighted_ than uniform DC (FDR-adjusted *P*=0.0062; two-sided Wilcoxon test; **Fig. 1D**). However, patients with mixed response showed a similar variation on RECIST_weighted_ *versus* uniform PD (*P*=0.27), and were significantly higher than uniform DC (*P*=0.021; **Fig. 1D**). Those results suggested that for patients with some levels of organ response, patients with a higher level of organ response heterogeneity are more likely to progress.

### Disease control and resistance to ICI or BRAF differ between and within metastatic sites

Tumor growth or reduction across cohorts was stratified by best response at the patient level comparing DC/PD. Among all ICI experiencing DC, liver (*n*=46) and lung (*n*=92) metastases experienced greatest reduction in tumor (−66%±8% and −63%±5%, respectively; mean±S.E.M.; **Fig. S3A**; I+N and PD1 each are shown in **Fig. S3B** and **S3C**). For BRAF, liver metastases experienced the greatest reduction (−58%±8%) (**Fig. S3D**). Additional tumor volume metrics are provided in **Table S4**.

To assess intra-organ heterogeneity of response to ICI, tumor volume changes are reported in patients with multiple lesions in the same organ. For patients who received ICI with multiple same-organ target lesions, brain metastases demonstrated the highest variability in intra-organ response comparing RECIST_weighted_ of lesions 01 *versus* 02 (FDR-adjusted *P*=0.084, two-sided Wilcoxon test for paired samples) (**Fig. 1E**). For all organ sites, the largest lesion at baseline was defined as 01 and second as 02. No substantial inter-lesion heterogeneity was observed across metastatic sites for BRAF. In addition, the absolute differences in the two lesions were computed and compared between overall response groups. Lung metastases had higher intra-organ absolute differences in DC/PD (FDR-adjusted *P*=0.074; two-sided Wilcoxon test; **Fig. S4**). Adding clinical variables, we observed that non-cutaneous melanoma showed significantly higher RECIST_weighted_ than cutaneous melanoma in liver metastases (**Fig. S5**).

### Radiomic signature differentiates disease control rate at the patient and organ levels after BRAF targeted and immunotherapy

To identify radiomic features associated with DC or PD, we performed comparisons between DC/PD at the patient level, or within each organ. Starting with I+N, we detected 39 differential radiomic features at FDR 0.10 (**Fig. 2A**). Of these, we found that the DC/PD differences were mostly driven by lung compared with other organ sites (**Fig. S6**) however this was not shared by PD1 or BRAF.

**Figure 2.**
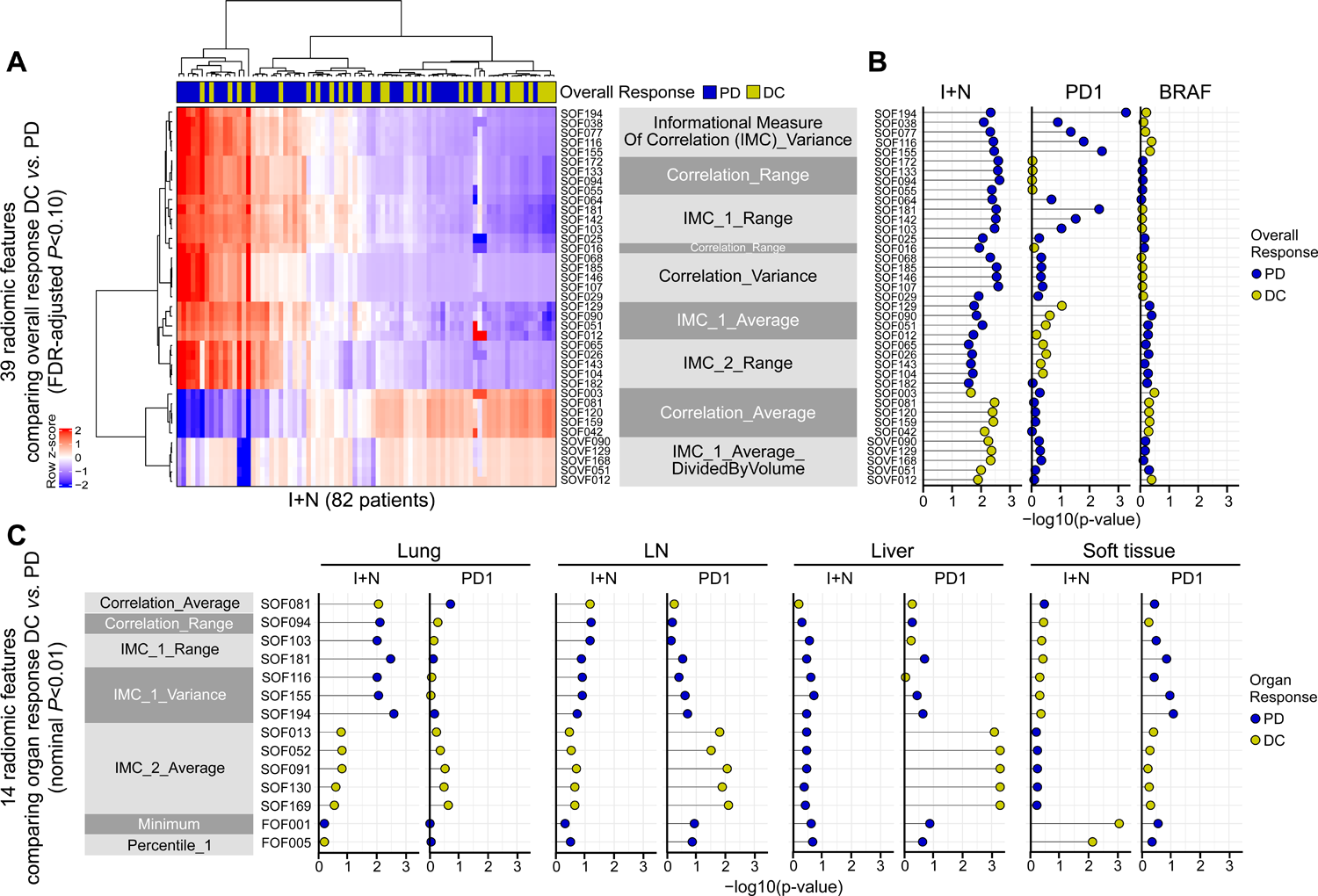
Radiomic features differentiate overall response or organ-specific response DC *versus* PD. (**A**) 39 features that distinguish overall response in patients who received I+N at FDR-adjusted *P*<0.10. Patients are clustered on the column and features are clustered on the row with dendrograms shown. The horizontal annotation bar on top of the heatmap indicates overall response PD and DC. Feature names (e.g., IMC_1_Variance) are shown on the right side of the heatmap, which correspond to the feature IDs (e.g., SOF194, SOF038, etc.). *n*=82 patients from I+N cohort are shown. (**B**) Overlapping or unique features across patient cohorts. The DC *versus* PD differences of the 39 features from **A** (I+N) are shown in patients who received PD1 monotherapy or BRAF targeted therapy. Features are shown in the same order as on the heatmap from **A**. (**C**) 14 features that distinguish organ-specific response in one or more cohorts. (left to right) each organ site is shown in I+N or PD1: lung, LN, liver, and soft tissue. LN = lymph node. IMC = Informational Measure of Correlation. For **A** and **C**, full feature IDs and names are described in **Tables S2** and **S3**. Wilcoxon rank-sum test was used in **A**, **B**, and **C**. All tests are two-sided.

Across the three cohorts, the I+N features were partially detected comparing DC with PD in the PD1 cohort, but absent in BRAF (**Fig. 2B**). Upon comparing DC/PD within each organ, we found that I+N and PD1 showed non-overlapping features associated with organ level response (**Fig. 2C**; nominal *P*<0.01). In lung metastases, seven radiomic features of “correlation average”, “correlation range”, “informational measure of correlation [IMC]_1 range”, and “IMC_1 variance” are associated with outcome in I+N, which were not detected in PD1 (**Fig. 2C, left**). At LN and liver metastases, I+N cohort did not return significant features, while PD1 cohort showed differences in “IMC_2 average” (**Fig. 2C, middle**). In soft tissue metastases, two features “minimum” and “percentile_1” which are first-order radiomics features distinguish DC from PD organs in I+N cohort, and no differences were detected in PD1 (**Fig. 2C, right**). Collectively, our results suggested that binary group comparisons distinguish radiomics features in I+N cohort with organ heterogeneity, and organ-specific features were specific to I+N or PD1 cohort each.

### Radiomics features predict response at the patient level and integrating clinical features improves the performance of DC *versus* PD classification

To predict overall response across all metastatic sites per individual, we evaluated 267 patients with high-quality images and performed radiomics modeling (221 ICI; 86 I+N & 135 PD1). We constructed ML models classifying DC/PD within I+N and PD1 each using XGBoost by 80/20 training/test with 10-fold cross-validation (CV) for feature selection and model optimization. Training/test set samples were randomly split prior to any processing steps, ensuring that samples in the test set were blind from the training set. In the training set, after proper scaling and normalization, features that are highly correlated (Spearman’s correlation ρ > 0.80), show high collinearity, or low variance were removed before model construction. This step reduced the number of radiomic features from 400 to approximately 10-30 total, which were then used for feature selection and hyperparameter tuning via 10-fold CV in the training set. The performance of the final optimized model on unseen data is reported in the test set.

The training CV AUC and test AUC are shown in **Fig. 3**. XGBoost models consisting of optimized radiomic signatures classified DC from PD across I+N (AUC = 0.71, CV; 0.85, test set; **Fig. 3A**, **Fig.S7A**), PD1 (AUC = 0.65, CV; 0.71, test set; **Fig. 3B**, **Fig.S7B**). Integration of clinical variables (age, body mass index (BMI), sex, AJCC stage, melanoma subtype, baseline eosinophil count, LDH, and NLR) to the radiomic signature achieved an improved predictive model for I+N (AUC = 0.71, CV; 0.89, test set; **Fig. 3C**), PD1 (AUC = 0.70, CV; 0.90, test set; **Fig. 3D**). We found that BMI, albumin, age, and eosinophil count were important predictors for I+N (VarImp ≥20; **Fig. 3E**). These four, plus NLR and AJCC M1c stage, also ranked as the top variables for PD1 model (**Fig. 3F**).

**Figure 3.**
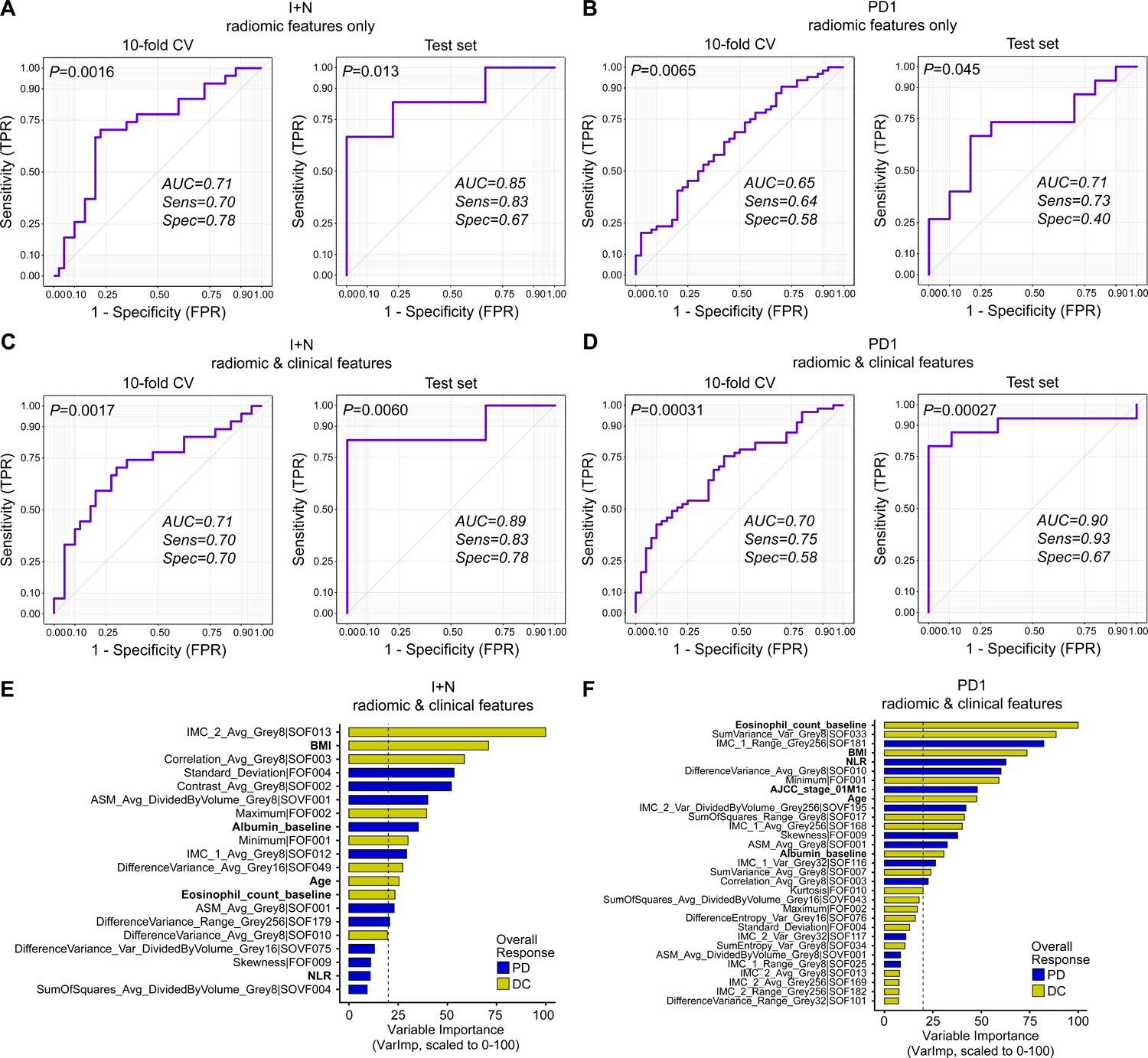
Radiomics models predict overall response DC/PD in ICI cohorts. For each cohort, models were optimized in the training set with 10-fold CV, and the final performance was reported on unseen data in the test set. We show both the training set 10-fold CV ROC curve as well as the test set ROC curve. AUC, Sensitivity (Sens), and Specificity (Spec) were reported. (**A**) Model of radiomic features only in I+N cohort. (**B**) Model of radiomic features only in PD1 cohort. (**C**) Model of radiomic features and clinical variables in I+N cohort. (**D**) Model of radiomic features and clinical variables in PD1 cohort. For I+N models in **A** and **C**: *n*=67 and 15 patients in training/test set (80% / 20% split), respectively (total is 82). 400 radiomic features were reduced to 17 prior to model training. For PD1 models in **B** and **D**: *n*=104 and 25 patients in training/test set (80% / 20% split), respectively (total is 129). 400 radiomic features were reduced to 23 prior to model training. (**E**) Variable importance (VarImp) of the features from I+N model in **C**. (**F**) Variable importance (VarImp) of the features from PD1 model in **D**. Features with VarImp >1 are shown in **E** and **F**; red vertical dashed line indicates VarImp=20; features with VarImp ≥20 are generally considered important in predicting outcome. Color indicates whether a feature is greater in overall response PD (blue) or DC (gold). Clinical variables are bolded. ROC = Receiver Operating Characteristic. AUC = Area Under Curve. CV = cross-validation. FPR = false positive rate. TPR = true positive rate. IMC = Informational Measure of Correlation. ASM = Angular Second Moment. The AUC p-value shown at the top left corner of each ROC panel in **A-D** was computed using function *roc.area* from R package verification (v1.42), which implements a two-sided Wilcoxon rank-sum test.

### Radiomics features predictive of organ-specific response vary by site of metastasis in ICI cohorts

We constructed radiomics models to predict response at metastasis level for I+N or PD1. To maximize the number of samples for model construction, we included all samples for training with LOOCV. Features were selected based on a bootstrapping strategy using 80% of the samples, repeated 100 times. In each bootstrapped set, we compared features between DC/PD within each organ and selected the top 10 features that consistently passed a lenient p-value threshold (nominal *P*<0.20) across bootstrapped sets (see **Methods**). The performance of the final model was reported based on LOOCV from the training set, recognizing that the generalization of the models will require independent validation. From I+N, the best performance of organ-specific models was observed in LN (AUC = 0.85; **Fig. 4A**, **Fig. S8A**). For PD1, the best performance of organ-specific models was observed in liver (0.94; **Fig. 4B**, **Fig. S8B**).

**Figure 4.**
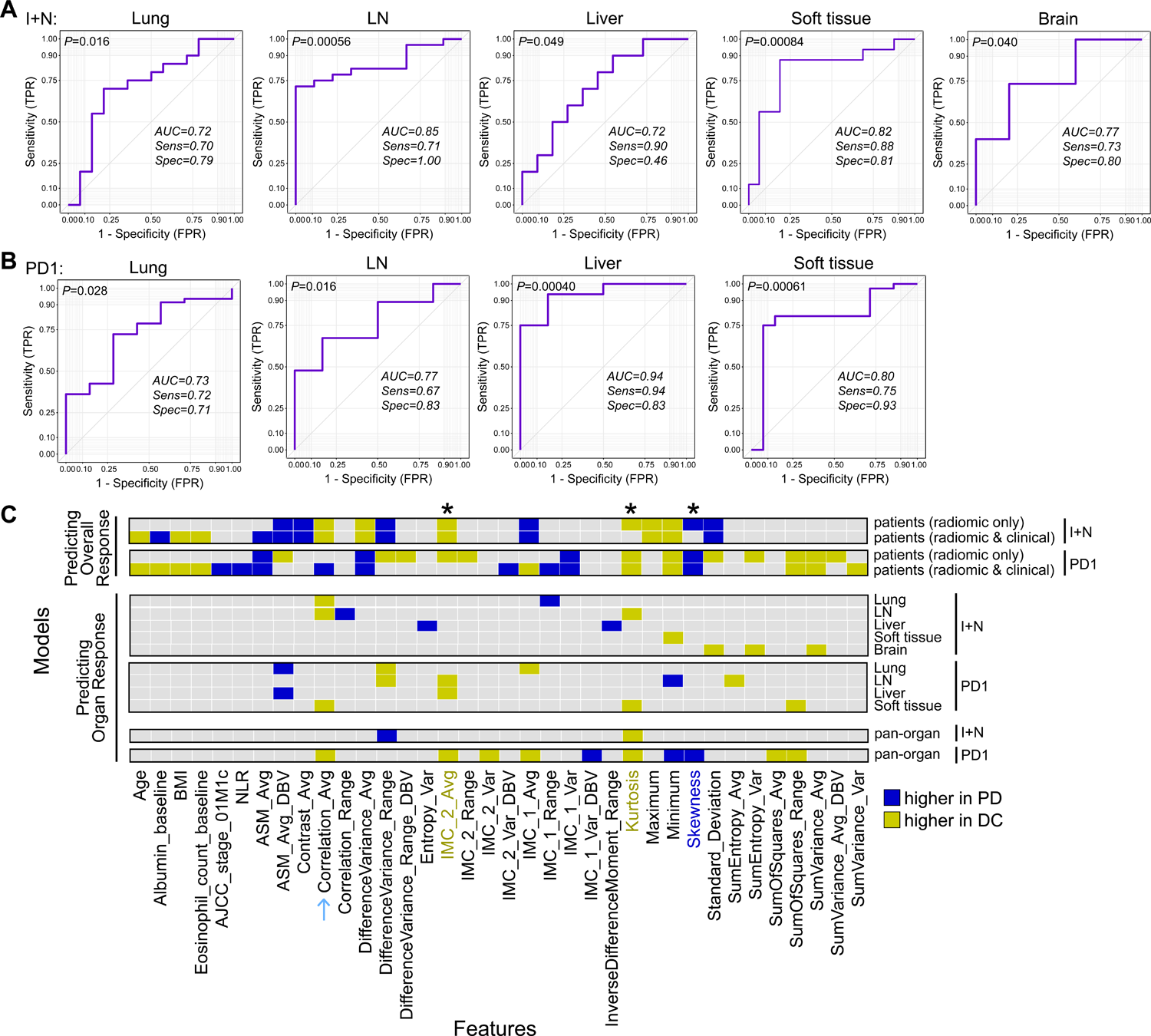
Radiomics models predict organ-specific response DC/PD in ICI cohorts. For each cohort, LOOCV was applied to all samples to generate the ROC curve. AUC, Sensitivity (Sens), and Specificity (Spec) were reported. 400 radiomic features were reduced to 10 prior to model construction. (**A**) Model of radiomic features in I+N cohort. (left to right): lung (34), LN (37), liver (21), soft tissue (32), and brain (20). Numbers in paratheses indicate the number of metastases for model construction per organ. (**B**) Model of radiomic features in PD1 cohort. (left to right): lung (54), LN (52), liver (22), soft tissue (50). Brain models of PD1 cohort were not constructed considering the small sample size. (**C**) Heatmap summarizing radiomic features utilized by overall response or organ-specific response models to predict DC/PD. Models are shown on the row, and features are shown on the column. Same radiomic features at different grey levels were collapsed as one entry for visualization purposes. Features of variable importance (VarImp) ≥ 20 from each model were included in these comparisons. Asterisks highlight top shared features high in DC (IMC_2_Avg, Kurtosis) or high in PD (Skewness). Blue arrow indicates features shared by I+N organ models (Correlation_Avg). LN = lymph node. ROC = Receiver Operating Characteristic. AUC = Area Under Curve. CV = cross-validation. FPR = false positive rate. TPR = true positive rate. IMC = Informational Measure of Correlation. ASM = Angular Second Moment. DBV = divided by volume (indicating this is a volume-independent second-order feature). The AUC p-value shown at the top left corner of each ROC panel in **A** and **B** was computed using function *roc.area* from R package verification (v1.42), which implements a two-sided Wilcoxon rank-sum test.

The radiomic features of each model were compared across cohorts or metastatic sites to assess features common or unique to each organ site. Same features at different grey levels were collapsed as one. Across all models, “kurtosis” and “IMC_2 average” are the most shared DC-related features, whereas the most shared PD-related feature is “skewness” (**Fig. 4C, asterisks**). For the patient-level models, I+N radiomics only and radiomics+clinical models shared ∼60% of features, while PD1 radiomics only and radiomics+clinical models shared ∼30% of features (**Fig. 4C, upper panels**). Looking into models constructed at each organ site, the majority of the features are unique to each organ, with overlapping ones including “kurtosis” associated with DC, among others (**Fig. 4C, middle panels**). When comparing models for I+N, “correlation average” was a shared feature across the lung and lymph node models (**Fig. 4C, blue arrow**). Taking into account different organ sites, we sought to build a pan-organ model that includes all metastases in predicting organ response DC/PD, with organ site as a covariate (lung, LN, etc.). We confirmed that individual patients’ organ metastases were either all in training or test set to prevent data leaking. The pan-organ models reached an AUC of 0.63 (CV) and 0.75 (test set) in I+N, 0.68 (CV) and 0.79 (test set) in PD1 (**Fig. S10**). **Table S5** describes the full list of feature comparisons between models.

## Discussion

Utilizing CT and MRI images from patients with melanoma treated with anti-PD1, I+N, and BRAF therapy, we have described organ-specific patterns of DC/PD and have built clinically informed radiomic models that may identify sites of likely progression. We observed that hepatic metastases experienced a significant reduction in size following ICI in DC but also the greatest increase in tumor volume in PD. Consistent with the literature that response and survival are attenuated in patients with liver metastases^24 25^, these data emphasize the liver as a primary driver of patient outcome in the context of ICI. In contrast, no variability in BRAF was observed. Few groups have produced models capable of predicting response at the metastatic lesion level, opting instead to average features from all lesions, or select a single representative lesion to analyze^26^. The ability to anticipate organ-specific resistance could potentially have clinical utility. Augmentation of systemic treatment regimens with localized therapy based on organ-specific resistance modeling could reserve treatments for subsequent therapy.

Using radiomic features derived from tumors at each metastatic site, we generated predictive models at both the patient and individual organ levels. With caution based on our sample size, we observed that our models performed well compared to published models, especially when including clinical variables^19^. We generally observed non-overlapping radiomic features driving model performance, potentially suggesting unique biology driving the anti-tumor immunity effect from each treatment, within each organ. Several radiomic features we identified have been associated with outcomes in cancer previously. Kurtosis, a measure describing the “peaked-ness” of the voxel intensity distribution of mass, is higher in homogenous lung nodules relative to heterogenous ones^27^ and associated with good prognosis in metastatic melanoma^28^. Consistent with this, a higher baseline kurtosis distinguishes DC from PD in our study at both the patient and organ levels (LN). The same pattern was observed in IMC_2, a gray-level co-occurrence matrix (GLCM) statistic that quantifies the complexity of texture and has been reported to be associated with clinical benefit in NSCLC^29^ and gastroenteropancreatic neuroendocrine tumours^30^. Other features identified (e.g., entropy) describe tumor spatial heterogeneity associated with inferior clinical outcome^31–33^. It may be worth noting that radiomic heterogeneity has been described to predict outcome to a similar degree to that of tumor size across cancer types^26 34^.

We emphasize the need for further validation in larger sample sets. Based on post-treatment “real-world” identification, we cannot control for lines of treatment, number and distribution of metastases, or other clinical factors. Our data were aggregated from patients treated within a large health system and we therefore cannot account for differences that might be due to variation between radiology machines in generating scan images. We also acknowledge that the statistical power of our models is less robust given our sample sizes, though only a few studies have described impressively larger melanoma sample sizes.

In summary, we describe patterns of treatment response and resistance as well as clinically informed radiomic predictive models that can identify individual sites of treatment refractory lesions. With validation of this work, future translational investigation of resistance and potentially clinical trials could be enhanced through the integration of these approaches.

## Data Availability

Data relevant to this study is provided in supplementary tables. Other data will be provided upon request from the corresponding authors.

## Declarations

## Ethics approval and consent to participate

The study protocol was approved by The University of Pittsburgh Institutional Review Board (IRB)-approved protocol (Protocol No. STUDY20020107). Participants gave informed consent to participate in the study before taking part. All samples have written informed patient consent.

## Consent for publication

All authors consent.

## Authors’ contributions

R.B. and J.J.L. conceived and designed the overall study. R.B. oversaw computational data analysis and machine-learning work. J.J.L. oversaw clinical review, data collection, and curation. R.B. and J.J.L. conducted editorial oversight. Z.N.G. wrote IRB. Z.N.G. and A.T. designed methods for clinical data, identified patients from the registry, and performed clinical data annotation and analysis. R.E.D. performed lesion heterogeneity analysis and comparisons. S.Z. coordinated radiologic evaluation and performed RECIST measurements and lesion segmentation. N.B. assisted with RECIST measurements and lesion segmentation. S.N. coordinated data collection and processing. A.A. assisted with clinical data acquirement. M.A. and N.A. exported radiologic images. T.Y.P. assisted with exporting radiologic images and lesion segmentation. V.P. performed radiomic feature extraction. P.M. assisted with preliminary bioinformatic and statistical analysis. S.B. assisted with clinical data annotation. A.G. and D.L.P saw patients in the clinic. J.M.K., H.M.Z., Y.G.N., D.D. contributed tumor and patient data. R.C. performed image processing and textual feature extraction from scans. R.B. designed and implemented the computational work for the development of machine-learning models. Z.N.G., A.T., R.E.D., J.J.L., and R.B. wrote the manuscript. All authors contributed to the final manuscript.

## Acknowledgments

The authors thank F. Mu (U. Pittsburgh) for their technical assistance in software installation and job execution on the HPCs.

## List of abbreviations

AJCC: American Joint Committee on Cancer
ASM: AngularSecondMoment
AUC: area under the ROC
BMI: body mass index
CR: complete response
CV: cross-validation
DBV: divided by volume
DC: disease control
FDR: false discovery rate
FOF: frist-order feature
GLCM: gray level co-occurrence matrix
I+N: nivolumab plus ipilimumab
ICI: and immune-checkpoint inhibitors
IMC: informational measure of correlation
LDH: lactate dehydrogenase
LOOCV: leave-one-out cross-validation
MEL: melanoma
NLR: neutrophil-to-lymphocyte ratio
NSCLC: non-small cell lung cancer
PD: progressive disease
PR: partial response
RECIST: Response Evaluation Criteria in Solid Tumors
ROC: Receiver Operating Characteristic
SD: stable disease
SEM: standard error of the mean
SOF: second-order feature
SOVF: second-order feature divided by volume
ULN: upper limit normal
VarImp: variable importance.

