## Supplementary material for "Radiomic analysis of patient and inter-organ heterogeneity in response to immunotherapies and BRAF targeted therapy in metastatic melanoma": Supp methods & Supp Fig1-9

### For

**Running Title:** melanoma organ-specific radiomics

**Feature**: Immunotherapy Biomarkers


**Authors**: Alexandra Tompkins[1,2*], Zane N. Gray[1,2*], Rebekah E. Dadey[1,2], Serafettin Zenkin[1,3], Nasim Batavani[1,3], Sarah Newman[1,2], Afsaneh Amouzegar[1,2], Murat Ak[1,3], Nursima Ak[1,3], Taha Yasin Pak[1,3], Vishal Peddagangireddy[1,3], Priyadarshini Mamindla[1,3], Sarah Behr[1], Amy Goodman[1], Darcy L. Ploucha[1], John M. Kirkwood[1,2], Hassane M. Zarour[1,4], Yana G. Najjar[1,2], Diwakar Davar[1,2], Rivka Colen[1,3], Jason J. Luke[1,2#], Riyue Bao[1,2#]

[1]UPMC Hillman Cancer Center, Pittsburgh, PA
[2]University of Pittsburgh, Department of Medicine, Pittsburgh, PA
[3]University of Pittsburgh Department of Radiology, Pittsburgh, PA
[4]University of Pittsburgh, Department of Immunology, Pittsburgh, PA
*Equal Contribution.
[#]Co-senior and corresponding authors.

[#]**Corresponding Authors:**
Jason J. Luke, MD, FACP
Associate Professor of Medicine
UPMC Hillman Cancer Center
5150 Centre Ave. Room 1.27C
Pittsburgh, PA 15232


Riyue Bao, PhD
Associate Professor of Medicine
UPMC Hillman Cancer Center
5150 Centre Ave, Suite 1A, Room 105
Pittsburgh, PA 15232




**Keywords:** immunotherapy, checkpoint inhibitors, radiomics, machine learning, response prediction, organ-specific, metastatic, melanoma

This document contains the full description of **Methods** for the manuscript, and **Supplementary Figures 1 to 9. Supplementary Tables 1 to 5** are provided in a separate spreadsheet.

### Supplementary Methods

#### Patient populations

The UPMC Hillman Cancer Center registry was queried for unresectable stage III/IV melanoma from 2015-2020, who also received anti-PD1 (pembrolizumab or nivolumab; PD1 cohort), anti-PD1+anti-CTLA4 (nivolumab plus ipilimumab; I+N cohort), or BRAF±MEK inhibitors (vemurafenib, dabrafenib, encorafenib ± cobimetinib, trametinib, binimetinib; BRAF cohort) treatment (adjuvant therapy were excluded) (**Table S1**). Patients treated with ipilimumab before pembrolizumab or nivolumab were excluded given evidence that anti-CTLA4 therapy impacts molecular characteristics of anti-PD1 response[1]. A detailed workflow of patient selection is illustrated in **Fig. S1**. The study protocol was approved by The University of Pittsburgh Institutional Review Board (IRB)-approved protocol (Protocol No. STUDY20020107). All samples have written informed patient consent.

#### Clinical data annotation and therapy response

Demographic, clinical, treatment, and radiologic variables measured using RECIST were collected for each patient from electronic medical record (EMR) databases as per our IRB-approved protocol (**Table S1**). For each cohort (PD1, I+N, or BRAF), we aggregated patients across lines of therapy based on the assumption that radiomic features of treatment response/resistance would be similar[2]. For each patient, tumor sizes and best overall response were evaluated by a board-certified radiologist (S.Z., N.B.) by Response Evaluation Criteria in Solid Tumors (RECIST) v1.1[3]. Clinical outcomes were categorized into two groups: disease control (DC; including complete response [CR], partial response [PR], stable disease [SD]) and progressive disease (PD). Patients with high-quality CT and/or MRI imaging with measurable disease available were included in radiomics analysis (**Fig. S1**). A maximum of two lesions per organ were measured following RECIST v1.1 guidelines[3]. An illustration of the full analysis workflow is provided in **Fig. S2**.

##### Organ-level weighted RECIST scores

Given that radiomics data were generated from scans of target lesions, we computed a weighted RECIST score at the organ level to directly link radiomics with response derived from target lesions. The formula of $RECIST_{weighted}$ is as follows:

(1) if there is only one target lesion at an organ site, $RECIST_{weighted} =$

$$\frac{lesion\ size\ at\ best\ response - lesion\ size\ at\ baseline}{lesion\ size\ at\ baseline},$$

(2) if there are two target lesions at an organ site, $RECIST_{weighted} =$

$$\frac{(lesion\ 01\ size\ at\ best\ response - lesion\ 01\ size\ at\ baseline) + (lesion\ 02\ size\ at\ best\ response - lesion\ 02\ size\ at\ baseline)}{lesion\ 01\ size\ at\ baseline + lesion\ 02\ size\ at\ baseline}$$

Subsequently, organ response categories were defined as PD if $RECIST_{weighted} \geq 20\%$, and DC if $RECIST_{weighted} < 20\%$.

##### Inter-organ heterogeneity analysis

To assess inter-organ heterogeneity, we computed the standard deviation (SD) of $RECIST_{weighted}$ scores across all organ sites per patient, and compared between patients with uniform progression, mixed response, and uniform DC using two-sided Wilcoxon rank-sum tests. One metastasis refers to all target lesions at the same organ site in the same patient. Tumors at different organ sites are considered different metastases. Only patients with two or more metastases were included in the analysis.

##### Intra-organ heterogeneity analysis

Within each organ site, a maximum of two lesions were measured. For all organ sites, the largest lesion at baseline was defined as 01 and second as 02. To assess intra-organ heterogeneity, we computed the tumor size change for lesions 01 and 02 separately as follows:

$$lesion\ 01\ tumor\ size\ change = \frac{lesion\ 01\ size\ at\ best\ response - lesion\ 01\ size\ at\ baseline}{lesion\ 01\ size\ at\ baseline}$$

$$lesion\ 02\ tumor\ size\ change = \frac{lesion\ 02\ size\ at\ best\ response - lesion\ 02\ size\ at\ baseline}{lesion\ 02\ size\ at\ baseline}$$

Subsequently, we compared tumor size changes of lesion 01 *versus* 02 within organ sites using two-sided Wilcoxon signed-rank tests. In addition, we computed the absolute value of the difference between lesion 01 and 02 tumor size changes $\Delta = |\ lesion\ 01\ tumor\ size\ change - lesion\ 02\ tumor\ size\ change\ |$, and compared the organ-specific Δ between the overall response groups PD and DC using two-sided Wilcoxon rank-sum tests after log10 transformation.

**Radiomics image processing and feature extraction**

Individual target lesions underwent semi-automatic segmentation with 3D Slicer[4] (v4.10.2) to create a volume of interest (VOI) for radiomics analysis from pretreatment contrast-enhanced CT scans and T1 pre-contrast, T1 post-contrast, and T2/FLAIR MRI sequences. Images were pre-processed using Nyul Intensity Normalization[5], a technique used to standardize the intensity levels of medical images, ensuring consistency and comparability across different scans and imaging modalities. It adjusts the pixel values within a scan, aligning them with a predefined intensity range acquired from a reference scan. Radiomic features were extracted from scans using Python (v3.8) packages including NumPy (v1.2) and SimpleITK (v2.2.0).

From each extracted VOI, a total of 400 features were derived, consisting of 10 histogram-based descriptors and 390 second-order texture features (**Table S2**). The description of the first-order and second-order features is provided in **Table S3**. The second-order features comprise 13 Haralick features[6], each computed across five gray levels and three rotation-invariant measures. The naming convention for second-order features is "(Gray-level Quantization)_(Feature Name)_(Measure used for Rotation Invariance)", where (Gray-level Quantization) denoted as “8,16,32,64,256”, (Feature Name) denoted as “Haralick feature name”, and (Measure used for Rotation Invariance) denoted as average (avg), variance (var), and range of the Grey-Level Co-occurrence Matrix (GLCM) features computed across four different angles to ensure their rotational invariance. Further details are described as follows.

The first-order features were computed to describe the distribution of voxel intensities within each Region of Interest (ROI). These included statistical metrics such as Minimum, Maximum, Mean, Standard Deviation, Percentiles (1, 5, 95, 99), Skewness, and Kurtosis.

The second-order texture features are derived from the GLCM, which characterizes the spatial relationship of pixel intensities within an image. GLCM captures the frequency of occurrence of pairs of pixel intensity values at given spatial displacements and directions, providing valuable information about texture patterns. Each second-order feature was derived from GLCMs computed with respect to four angles (0°, 45°, 90°, and 135°), representing the angles between the pixel of interest and its neighboring pixels, allowing for a comprehensive capture of variations from various perspectives.

To ensure rotation invariance, statistical measures including average, mean, and range were computed across all angles for each feature resulting in 60 rotation-invariant features. These measures effectively encapsulate the feature's essence, regardless of its orientation. To enhance the signal-to-noise ratio, the original images underwent discretization into five grey levels (8, 16, 32, 64, and 256 grey levels), with 60 rotation-invariant features computed at each grey level. Furthermore, the set of 195 features is further refined by dividing each feature by the VOI, resulting in another set of 195 volume-independent features, leaving 400 radiomics features for subsequent analysis.

For MR imaging, preprocessing steps were undertaken per Oxford Centre for Functional Magnetic Resonance Imaging of the Brain (FMRIB) Brain Extraction Tool (BET)[7] resulting in three distinct imaging phenotypes free of non-brain tissue: edema/invasion, active enhancing tumor and necrosis. Contralateral physiologic white matter was also segmented for within-sequence normalization[8] and hemorrhage, and intracranial vasculature appearing as hyperintensity on pre-contrast T1W1 images were subtracted from FLAIR sequence images to prevent obscuration of tumor textural features.

**Machine-learning model construction**

Machine learning (ML) to predict overall and organ-specific responses using radiomic features was carried out using eXtreme Gradient Boosting[9] (XGBoost). ML construction was performed using R package caret[10] (v6.0.91), with steps, functions, and parameters described below.

<u>To develop overall response DC/PD models</u>: We constructed XGBoost models to predict overall response in I+N cohort (*n*=82) or PD1 cohort (*n*=129). Data were randomly split into training and test sets by an 80/20 ratio using function *createDataPartition*. In the training set, features were assessed to

exclude the ones with low variance, high collinearity, or strong correlation (Spearman's correlation ρ > 0.8). This step reduced 400 radiomics features to 17 for I+N models and 23 for PD1 models. The selected features are listed in **Fig. S7**. Subsequently, model optimization and hyperparameter tuning (xgb_tune) were performed with 10-fold cross-validation (CV) using function *train*, with parameters [ $\mathrm{method} = \text{"xgbTree"}, \mathrm{trControl} = \mathrm{cv\_opts}, \mathrm{tuneGrid} = \mathrm{xgb.grid}, \mathrm{metric} = \text{"ROC"}$ ]. $cv\_opts$ is the CV object generated by function *trainControl* with parameters [ $\mathrm{method} = \text{"cv"}, \mathrm{summaryFunction} = \mathrm{twoClassSummary}, \mathrm{number} = 10, \mathrm{classProbs} = \mathrm{TRUE}$ ], and $\mathrm{xgb.grid}$ is the object generated by function *expand.grid* with parameters [ $\mathrm{nrounds} = 1000, \mathrm{eta} = \mathrm{c}(0.001, 0.005, 0.01, 0.02, 0.05, 0.1, 0.2, 0.3, 0.4, 0.5), \mathrm{max\_depth} = \mathrm{c}(2, 3, 4, 5, 6, 7, 8, 9, 10), \mathrm{gamma} = \mathrm{c}(1, 2, 3), \mathrm{subsample} = \mathrm{c}(0.5, 0.75, 1), \mathrm{min\_child\_weight} = \mathrm{c}(1, 2, 3), \mathrm{colsample\_bytree} = \mathrm{c}(1)$ ], representing various combinations of hyperparameters. The model of the highest AUC was selected as the final model. The model's performance on unseen data was evaluated in the test set using function *predict* with default parameters.

<u>To develop organ-specific DC/PD models</u>: We constructed XGBoost models to predict organ-specific response in I+N or PD1 cohort, including lung, lymph node [LN], liver, soft tissue (with CT features), and brain (with MRI features). The sample sizes varied across organs and cohorts: in I+N cohort, we had lung (*n*=34), LN (*n*=37), liver (*n*=21), soft tissue (*n*=32), and brain (*n*=20), while in PD1 cohort, we had lung (*n*=54), LN (*n*=52), liver (*n*=22), soft tissue (*n*=50). Due to the small sample size (<20) in the PD1 cohort for brain samples, we did not build models for brain responses in this cohort.

Recognizing that sample sizes for organ models are generally smaller than patient cohort sizes available for response prediction, and to ensure our methods were consistent for all organ models, we used a different approach from above. Data were not split into training and test sets; model's performance was evaluated through the Leave-One-Out CV (LOOCV) method, recognizing that the generalization of the models will require independent validation. For each organ site, a bootstrap strategy was implemented to reduce the initiated set of 400 radiomics features per organ to a more manageable number prior to model training. Taking lung from I+N cohort as an example, this process includes three steps:

(1) We initially selected features that were abundant in the majority of the samples. Specifically, we computed the arithmetic mean of each feature across 34 lung metastases, $feat\_mean(i)$ , where $i$ represents feature 1,2,3…400. Features with values higher than $feat\_mean(i)$ in at least 30% of the samples were retained, resulting a reduction from 400 features to 245;

(2) Subsequently, we performed bootstrapping using 80% of the samples. This was done by randomly sampling with replacement from the original dataset 100 times, creating 100 sets of samples. For each set of bootstrapped samples, we conducted two-sided Wilcoxon rank-sum tests to identify features differentially abundant between groups based on clinical outcomes (DC and PD). We kept features that passed $P<0.20$, which is a lenient threshold commonly used in the literature[11]. Adjustment for multiple comparisons was not applied in this scenario;

(3) Finally, we ranked the features based on their relevance to the outcome by summarizing the number of sample sets in which they were identified as differentially abundant, and selected top 10 features for model training. The selected features are listed in **Fig. S8**.

We repeated this procedure for all organs in I+N or PD1 cohort. The training process was performed using *train* function with [ $\text{method} = \text{"LOOCV"}$ ] as the *trainControl* parameter and other parameters as described above. The model of the highest AUC was selected as the final model.

<u>To develop pan-organ DC/PD models</u>: In addition to individual organ models, we constructed XGBoost models to predict pan-organ response across all metastases in I+N or PD1 cohort, with organ site included as a covariate. Data were split into training and test sets, with a procedure ensuring that metastases from the same patient fell into the same set (either training or test) to prevent data leakage. We treated metastases from the same patient as a single block, assigning each block a unique index. Patient blocks were then split by an 80/20 ratio using function *createDataPartition*, and the original organ-based training and test sets were reconstructed based on these indexes.

In the training set, features were assessed to exclude the ones with low variance, high collinearity, or strong correlation (Spearman's correlation $\rho > 0.8$). This step reduced 400 radiomics features to 15 for I+N models and 24 for PD1 models. The selected features are listed in **Fig. S9**. Model optimization and hyperparameter tuning were the same as that of overall response models described above. The model of the highest AUC was selected as the final model. The model's performance on unseen data was evaluated in the test set using function *predict* with default parameters.

**ROC generation and variable importance evaluation**

For overall response or pan-organ response DC/PD models, we reported two types of Receiver Operating Characteristic (ROC) curves for the final model. This includes: (1) the training set 10-fold CV ROC curve, representing the CV prediction by the final model during the training process. These ROC curves were generated using function *evalm* with parameters [ $\mathrm{list1} = \mathrm{xgb\_tune}, \mathrm{positive} = "\mathrm{DC}"$ ] from R package MLeval (v0.3). xgb_tune is the XGBoost training object generated by function *train* from R package caret (v6.0.91), with detailed parameters described in the model construction methods section above; and (2) the test set ROC curve, representing the performance of the final model on unseen data. These ROC curves were generated using function *roc* from R package pROC (v1.18.0). For organ-specific response DC/PD models, we reported the LOOCV ROC curve for the final model using function *evalm*.

For all models, sensitivity and specificity were also reported alongside AUC metrics. The p-values for AUC were computed using function *roc.area* from R package verification (v1.42), which implements a two-sided Wilcoxon rank-sum test to determine statistical significance. Variable importance (VarImp) of each model was computed using function *varImp* from R package caret (v6.0.91) with parameter [ $scale = TRUE$ ].

**Comparison of DC and PD groups for detection of differential radiomic features**

To independently evaluate the association between radiomics features and clinical outcomes, we performed statistical comparisons of DC and PD groups to directly detect differentially abundant radiomics features associated with the clinical outcome. Groups of at least five samples were included in this analysis. All data were first shifted to non-negative values by adding a constant, followed by log10 transformation. Starting with 400 radiomics features, we removed features that did not exceed their mean value in at least 30% of the samples prior to statistical analysis. Differentially abundant features were identified using two-sided Wilcoxon rank-sum tests. A FDR-adjusted p-value threshold of <0.10 was applied to comparisons between overall response categories (DC *versus* PD), and a nominal p-value threshold of <0.01 was applied to organ response comparisons.

**Statistical analysis**

Among the clinical variables, patients with high/low neutrophil to lymphocyte ratio (NLR) were stratified by NLR ≥ or < 3.0, and those with high/low lactate dehydrogenase (LDH) were stratified by LDH ≥ or < 170, per literature review for high-risk status and institutional laboratory standards for patients with metastatic melanoma[12 13]. The association between mixed organ responses in patients and PD as the overall response was assessed by Fisher's exact test. SD of metastasis site's $RECIST_{weighted}$ scores was compared between patients with uniform PD, mixed responses, and uniform DC by Wilcoxon rank-sum test. Metastasis site's $RECIST_{weighted}$ scores were compared between overall response groups DC/PD using Wilcoxon rank-sum test. The absolute differences in lesions' tumor size change% in lesion 01 *versus* 02 were log10-transformed and then compared between overall response groups DC/PD using Wilcoxon rank-sum test. The statistical significance of lesion's tumor size change% in lesion 01 *versus* 02 was determined by Wilcoxon signed-rank test. Metastasis site's $RECIST_{weighted}$ scores were compared between patients with cutaneous *versus* non-cutaneous melanoma using Wilcoxon rank-sum test. Differentially abundant radiomics features between DC and PD were identified using Wilcoxon rank-sum test. Other statistical tests were described in the relevant method sections above. FDR was controlled at 0.10. Benjamini-Hochberg (BH)-FDR procedure[14] was used for multiple comparison adjustments. All tests were two-sided.

#### Supplementary Figures

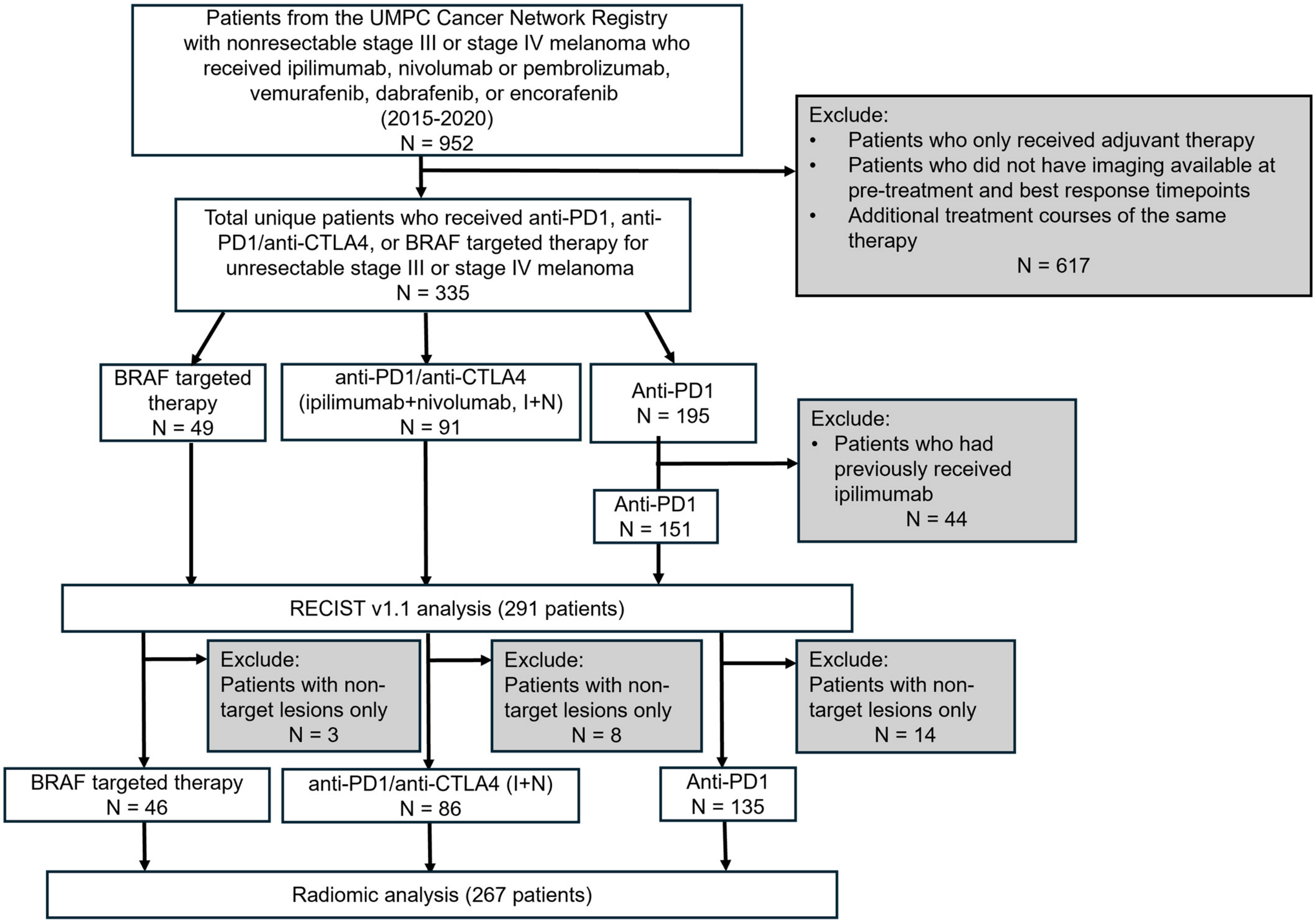


**Supplemental Figure 1. Workflow for identification of analysis population.** Patient selection process includes: (1) Identify patients who received ipilimumab/nivolumab (I+N), PD1 monotherapy, or BRAF targeted therapy and have appropriate imaging available; (2) Exclude patients who received therapy as an adjuvant treatment; (3) If a patient received the same therapy for multiple separate treatment courses only include in each cohort once, with a preference for earlier treatment course if appropriate imaging is available; and (4) In PD1 cohort exclude patients who had received ipilimumab prior to PD1 treatment.

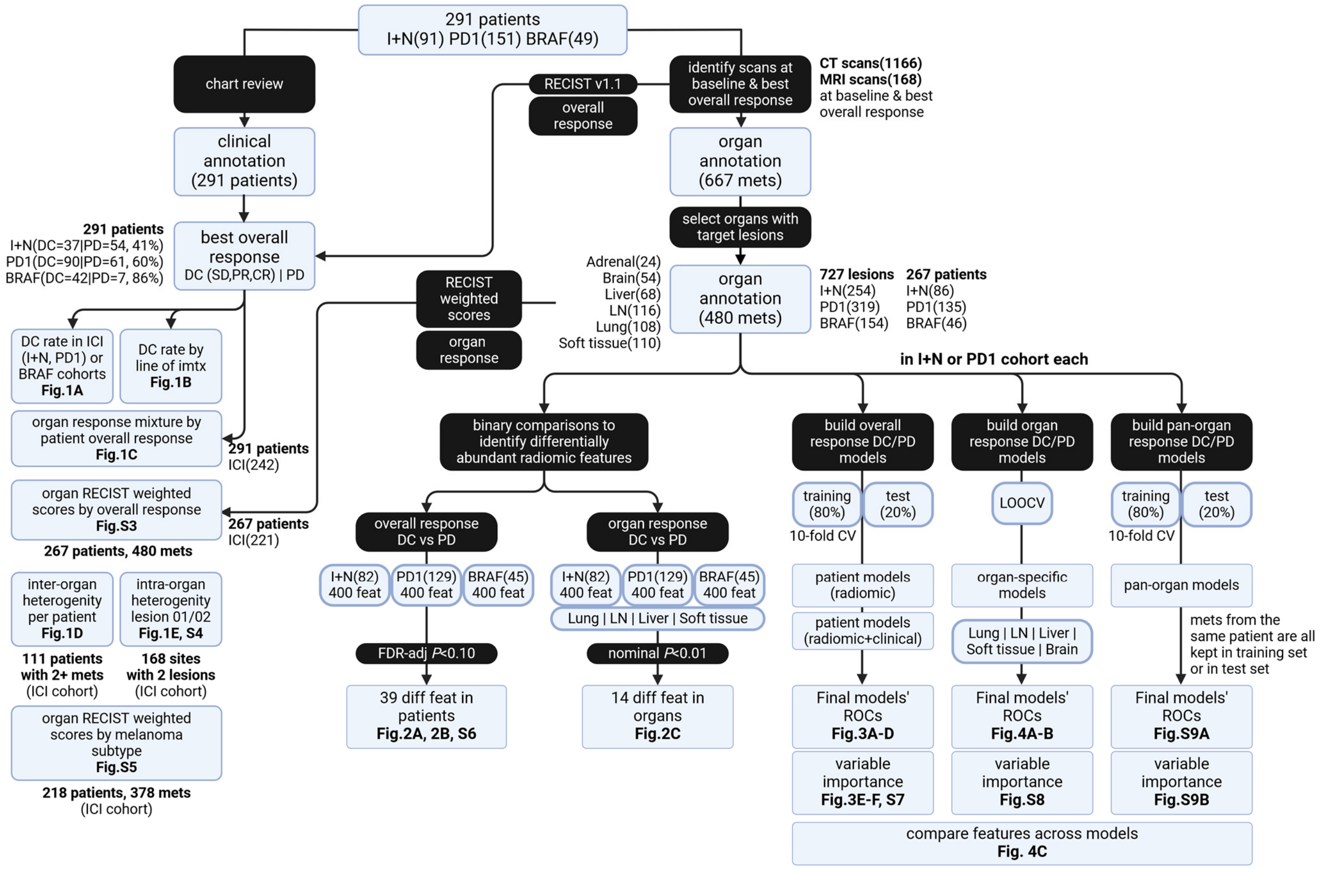


**Supplemental Figure 2. Overall analysis workflow.** Clinical data and medical images from 291 patients were used for the analysis of overall and organ-specific response, inter- and intra-organ heterogeneity, differentially abundant radiomics features in DC *versus* PD, and machine-learning models predicting overall response or organ-specific response. Multiple lesions at the same organ site are considered one metastasis. Mets = metastases. Imtx = immunotherapy. LN = lymph node. CV = cross-validation. LOOCV = leave-one-out CV. ROC = Receiver Operating Characteristic.

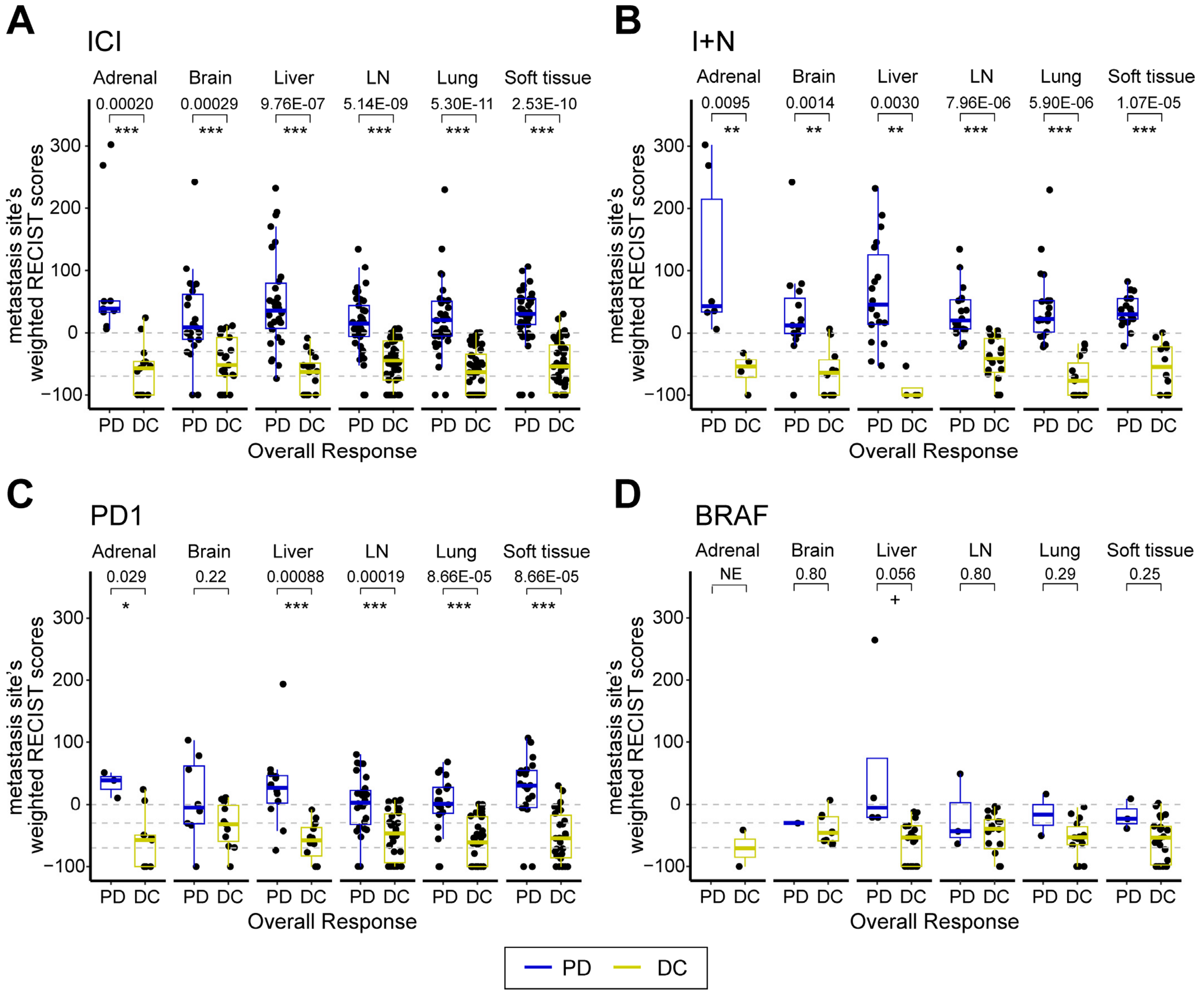


**Supplemental Figure 3. Organ's weighted RECIST scores comparing overall response differ by organ site.** (**A**) ICI cohort. (**B**) I+N cohort. (**C**) PD1 cohort. (**D**) BRAF cohort. Each data point represents one metastasis at each organ site. *n*=480 metastases from 267 patients are shown. LN = lymph node. Two-sided Wilcoxon rank-sum test was used in **A-D**. FDR-adjusted p-values are shown. FDR was controlled at 0.10. Denotations: *** $P$<0.001, ** $P$<0.01, * $P$<0.05, + $P$<0.10.

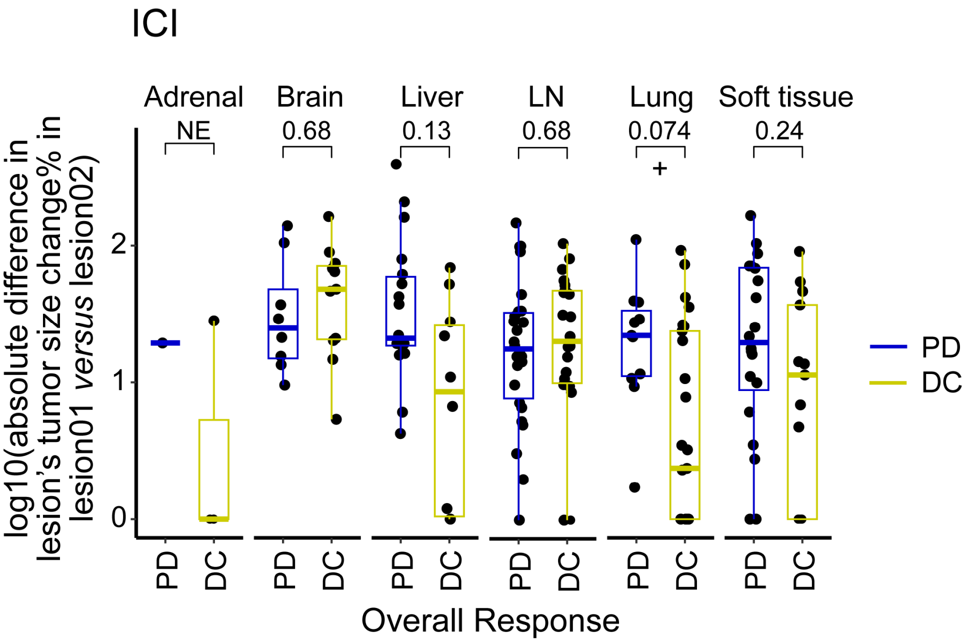


**Supplemental Figure 4. Intra-organ heterogeneity by overall response in ICI cohort.** The y-axis represents log10-transformed absolute differences in (lesion01's tumor size change% minus lesion 02's tumor size change%). *n*=168 sites from ICI patients who had two lesions per organ site are shown. LN = lymph node. Two-sided Wilcoxon rank-sum test was used. FDR-adjusted p-values are shown. FDR was controlled at 0.10. Denotations: + *P*<0.10.

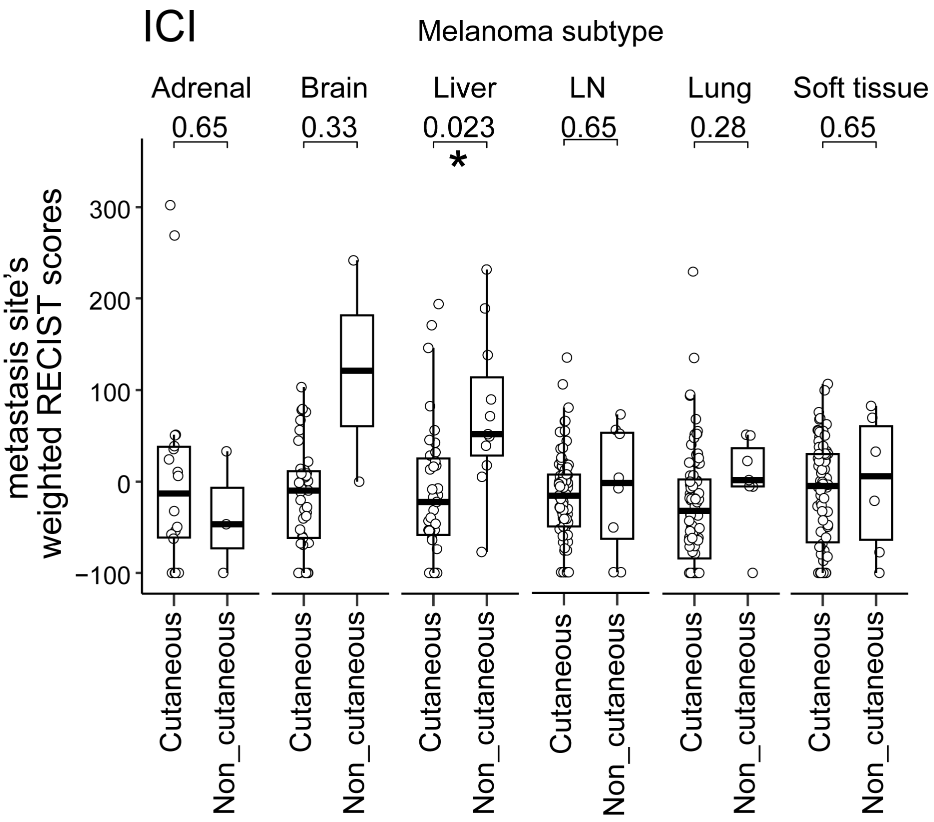


**Supplemental Figure 6. Organ's weighted RECIST scores by melanoma subtype in ICI cohort.** LN = lymph node. Two-sided Wilcoxon rank-sum test was used. FDR-adjusted p-values are shown. FDR was controlled at 0.10. Denotations: * *P*<0.05.

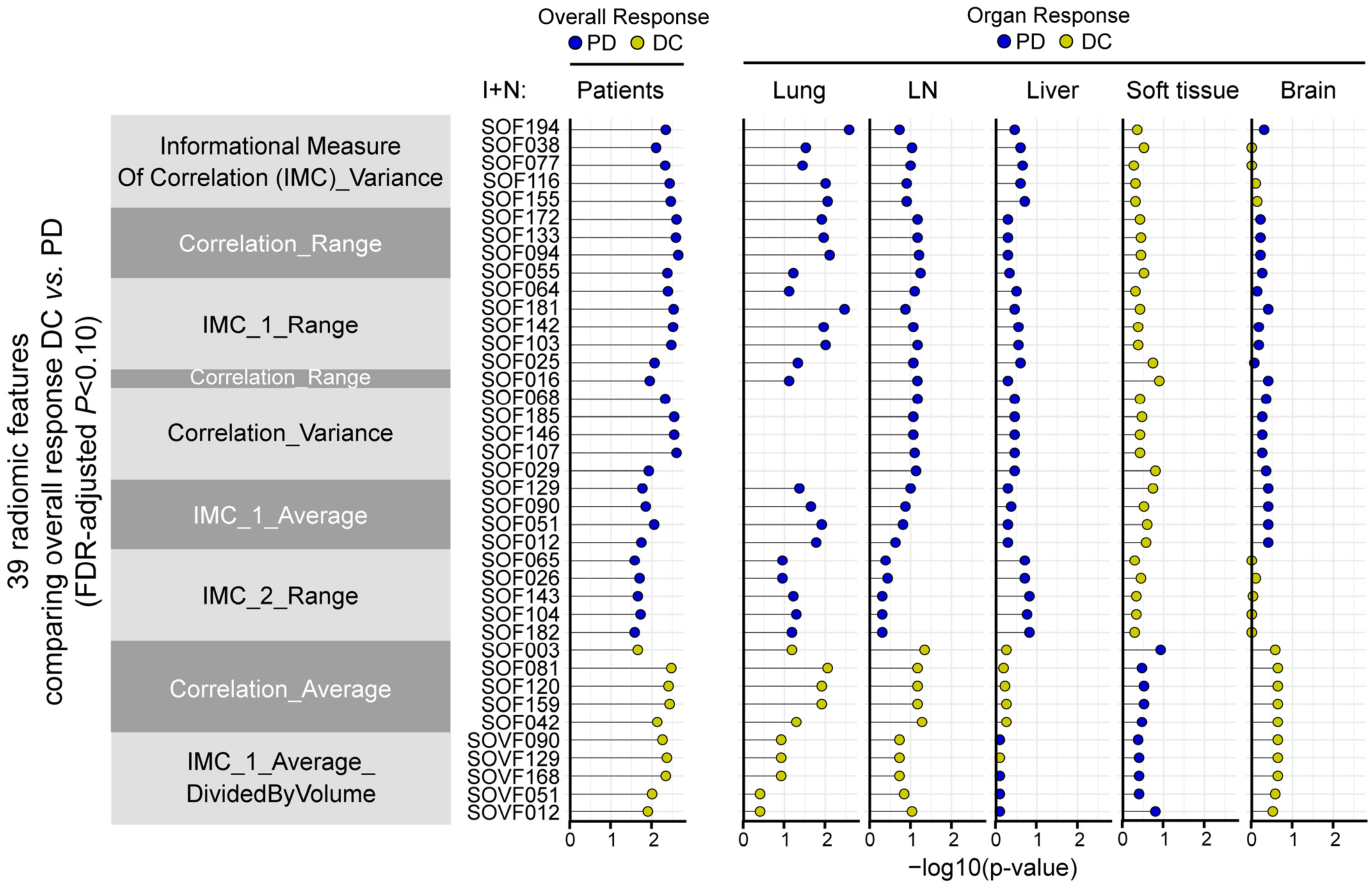


**Supplemental Figure 6. The distribution of the 39 radiomics features that distinguish overall response DC/PD in I+N cohort by organ site and with organ response colored by DC or PD.** Features are shown in the same order as on the heatmap from **Fig. 2A** (FDR-adjusted *P*<0.10). Color indicates whether a feature is greater in organ response PD (blue) or DC (gold). LN = lymph node. IMC = Informational Measure of Correlation. Full feature IDs and names are described in **Tables S2** and **S3**. Two-sided Wilcoxon rank-sum test was used.

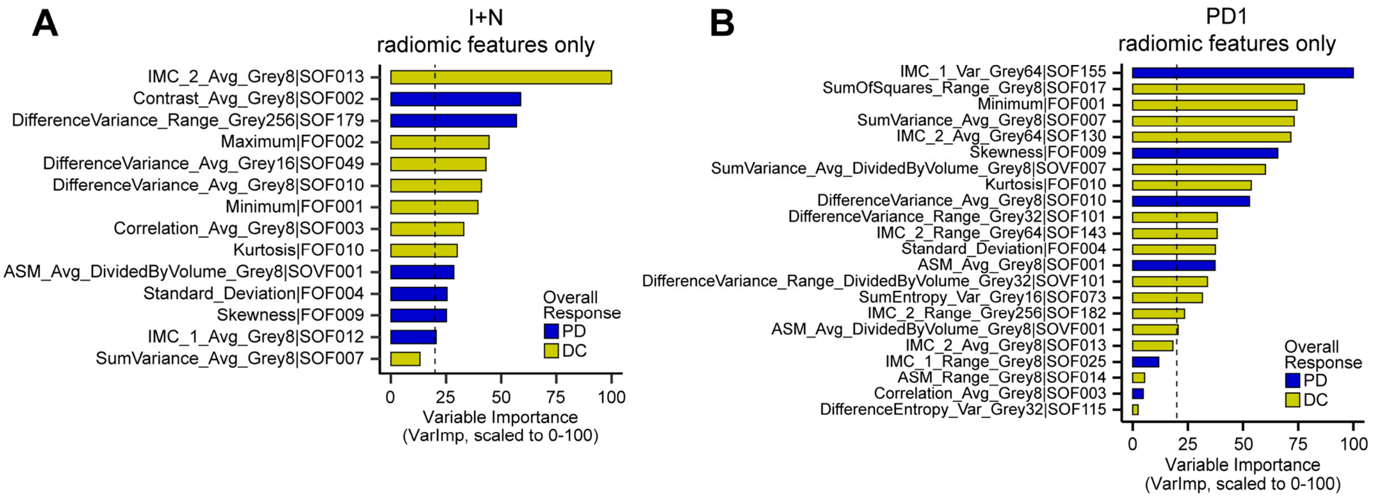


**Supplemental Figure 7. Variable importance (VarImp) of the radiomics only models predicting overall response DC/PD in I+N or PD1 cohort.** (**A**) I+N cohort, corresponding to the model in **Fig. 3A**. (**B**) PD1 cohort, corresponding to the model in **Fig. 3B**. Features with VarImp >1 are shown; red vertical dashed line indicates VarImp=20; features with VarImp ≥20 are generally considered important in predicting outcome. Color indicates whether a feature is greater in overall response PD (blue) or DC (gold). IMC = Informational Measure of Correlation. ASM = Angular Second Moment.

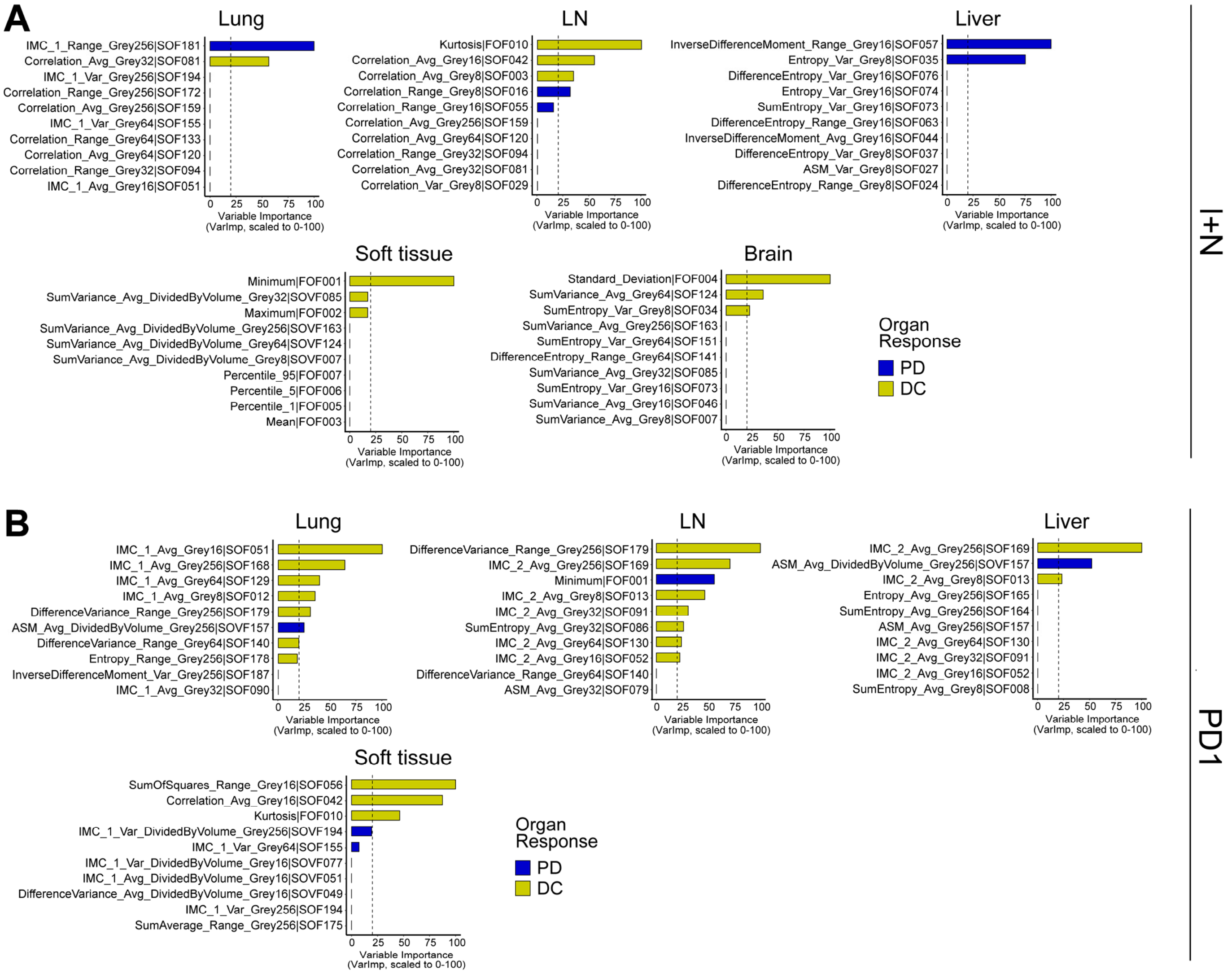


**Supplemental Figure 8. Variable importance of the radiomics models predicting organ-specific response DC/PD at each organ site from I+N or PD1 cohort.** (**A**) I+N cohort, corresponding to the models in **Fig. 4A**. (**B**) PD1 cohort, corresponding to the models in **Fig. 4B**. In each model, the 10 features used for model training are shown; red vertical dashed line indicates VarImp=20; features with VarImp ≥20 are generally considered important in predicting outcome. Color indicates whether a feature is greater in organ response PD (blue) or DC (gold). LN = lymph node. IMC = Informational Measure of Correlation. ASM = Angular Second Moment.

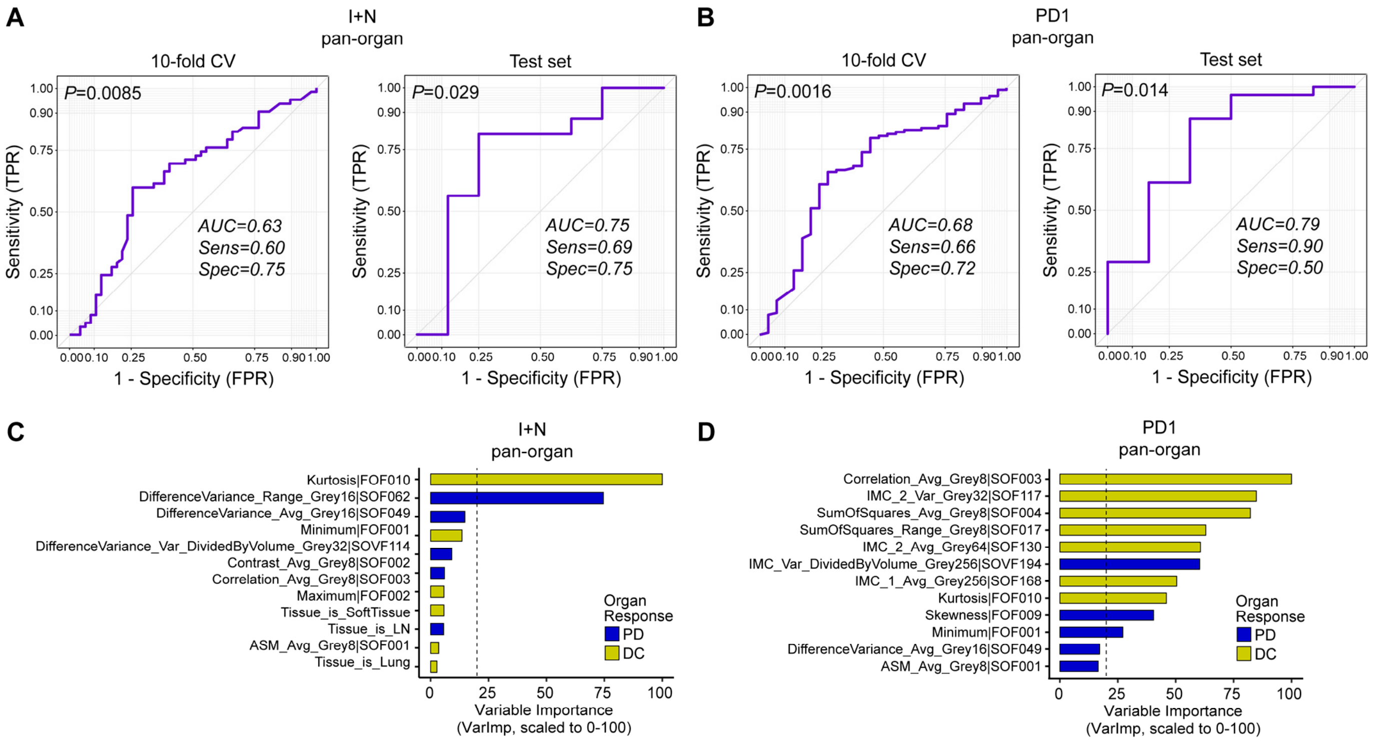


**Supplemental Figure 9. Pan-organ models predicting organ-specific response DC/PD in I+N or PD1 cohort.** Pan-organ models were developed across all metastases, with organ site as a covariate. We confirmed that individual patients' organ metastases were either all in training or test set to prevent data leaking. For each cohort, models were optimized in the training set with 10-fold CV, and the final performance was reported on unseen data in the test set. We show both the training set 10-fold CV ROC curve as well as the test set ROC curve. AUC, Sensitivity (Sens), and Specificity (Spec) were reported. (**A**) Model of radiomic features in I+N cohort. $n$=109 and 24 metastases in training/test set (80% / 20% split), respectively (total is 133). 400 radiomic features were reduced to 15 prior to model training. (**B**) Model of radiomic features in PD1 cohort. $n$=152 and 37 patients in training/test set (80% / 20% split), respectively (total is 189). 400 radiomic features were reduced to 24 prior to model training. (**C**) Variable importance (VarImp) of the features from I+N model in **A**. (**D**) Variable importance (VarImp) of the features from PD1 model in **B**. Features with VarImp >1 are shown in **C** and **D**; red vertical dashed line indicates VarImp=20; features with VarImp ≥20 are generally considered important in predicting outcome. Color indicates whether a feature is greater in organ response PD (blue) or DC (gold). ROC = Receiver Operating Characteristic. AUC = Area Under Curve. CV = cross-validation. FPR = false positive rate. TPR = true positive rate. IMC = Informational Measure of Correlation. ASM = Angular Second

Moment. The AUC p-value shown at the top left corner of each ROC panel in **A-B** was computed using function *roc.area* from R package verification (v1.42) which implements a two-sided Wilcoxon rank-sum test.

#### Supplementary Tables

**Supplemental Table 1. Demographic and clinical characteristics of the patients.**

**Supplemental Table 2. List of 400 radiomics features extracted from scans.**

**Supplemental Table 3. Description and definition of the radiomics features.**

**Supplemental Table 4. Tumor size changes per organ per cohort as shown in Fig. S3.**

**Supplemental Table 5. Comparison of radiomics features across models as shown in Fig. 4C.**
